# Association of Physical Activity with Change in Physical Function in Individuals with Atrial Fibrillation: The Atherosclerosis Risk in Communities (ARIC) Study

**DOI:** 10.64898/2026.08.18.26360678

**Authors:** Byung Joon Pae, B. Gwen Windham, Amit J. Shah, Linzi Li, Kathryn Wood, Elsayed Z. Soliman, Lin Yee Chen, Faye L. Norby, Amelia S. Wallace, Alvaro Alonso

## Abstract

**Background:** Atrial fibrillation (AF) is associated with declines in physical function. While physical activity is linked to better physical function in the general population, its long-term impact in people with AF remains unclear. Investigating this relationship could provide insights and inform interventions for this population.

**Methods:** 624 participants with AF from the Atherosclerosis Risk in Communities (ARIC) cohort assessed in 2011-2013 were studied. Physical activity was assessed using the modified Baecke Physical Activity Questionnaire. Physical function was measured using the Short Physical Performance Battery (SPPB), grip strength, and 4-meter walk time up to 3 times over an 8-year period, with 4-meter walk speed as a secondary outcome evaluated in supplemental analyses. Confounder-adjusted linear mixed models were used to assess associations between physical activity and change in physical function trajectories over time.

**Results:** Participants had a mean age of 78.5 ± 5.4 years, with 52.6% males and 13.8% Black. Median follow-up was 6.6 years. At baseline, greater sport-related leisure time, non-sport leisure time, and total moderate-to-vigorous physical activity (MVPA) were cross-sectionally associated with better physical function. However, physical activity measures were not significantly associated with temporal trajectories in physical function over time.

**Conclusions:** In participants with AF, greater habitual physical activity was significantly associated with better baseline physical function but not with future trajectories. Randomized trials are needed to examine whether interventions that improve habitual physical activity or MVPA can improve physical functioning in individuals with AF.

**Clinical Perspective:** *What is New?:*

- In this study, greater habitual physical activity was associated with higher SPPB, grip strength, and shorter 4-meter walk time in participants with AF at baseline.
- This study further found that greater habitual physical activity was not significantly associated with 6-year changes in SPPB, grip strength, and 4-meter walk time in participants with AF.

*What are the Clinical Implications?:*

- The findings imply that greater habitual physical activity is associated with better physical function in older adults with AF, but it may not be significantly associated with future functional trajectories.
- Future randomized intervention studies are required to determine whether structured exercise programs or higher-intensity physical activity can improve physical functioning in older adults with AF.

## Introduction

Atrial fibrillation (AF) is the most common clinically relevant arrhythmia, with a steady increase in global prevalence and incidence.^1, 2^ Estimates from the Global Burden of Disease study indicate that the worldwide prevalence of atrial fibrillation (AF) increased from 46.3 million to 59 million between 2016 and 2019.^3, 4^ Other evidence suggests a 4-fold rise in age-standardized prevalence over the past five decades.^5^ Given that AF is frequently undetected until symptoms occur, these figures likely underestimate its true burden.^1, 6^ AF, particularly in older adults, is associated with greater declines in physical function, including grip strength and gait speed, compared with individuals without AF.^7, 8^ Given the impact of these functional measures on quality of life and the risk of adverse clinical outcomes such as incident disability, institutionalization, and mortality,^9, 10, 11, 12^ there is a critical need for interventions that prevent or mitigate physical function decline in patients with AF.

Physical activity is associated with physical function,^13^ though the relationship between physical activity and physical function in patients with AF remains understudied. Prior studies have found beneficial effects of physical activity on physical function in patients with AF;^14^ however, most were limited by small sample sizes, assessment of short-term effects of exercise interventions. It remains unclear whether habitual physical activity in everyday life confers similar benefits for physical function among AF patients. Thus, larger studies assessing the long-term associations of habitual physical activity with physical function in patients with AF are needed. Such investigation could potentially provide additional insights regarding the association between physical activity and physical function in patients with AF and inform interventions that improve outcomes in this group.

This study aims to investigate the association between various measures of habitual physical activity and change in physical function in AF participants. We hypothesize that higher levels of habitual physical activity will be associated with slower declines in Short Physical Performance Battery (SPPB), grip strength, and 4-meter walk speed, as well as slower increases in 4-meter walk time among participants with AF.

## Methods

### Study Participants

Data was obtained from the Atherosclerosis Risk in Communities Study (ARIC), an ongoing community-based prospective cohort study established to investigate risk factors for cardiovascular disease (CVD).^15^ The study enrolled 15,792 adults aged 45–64 years between 1987 and 1989 (Visit 1) from 4 U.S. field centers: Washington County, MD; Forsyth County, NC; Jackson, MS; and suburbs of Minneapolis, MN. Participants subsequently attended follow-up examinations at Visit 2 (1990–1992), Visit 3 (1993–1995), and Visit 4 (1996–1998), with continued surveillance for clinical and functional outcomes. Visit 5 (2011–2013), which included 6,538 participants, served as the baseline for this study; Visit 6 (2016–2017) and Visit 7 (2018– 2019) served as follow-up assessments. Due to small sample sizes, (1) non-White participants from the Washington County and Minneapolis field centers and (2) participants who were neither White nor Black from the Forsyth County field center were excluded. Participants without a history of AF at or prior to baseline were further excluded. History of AF at or prior to baseline was determined by electrocardiograms and hospital discharge codes (ICD-9-CM: 427.3x). A total of 624 participants with a history of AF at or prior to baseline formed the final analytic sample for the primary analysis, in which multiple imputation was used to address potential bias due to missing data. For the complete case sensitivity analysis, 465 participants with a history of AF at or prior to baseline and without any missing baseline physical activity, physical function, and covariate data were included (Figure 1). Of the 465 participants, those with stroke (n=44), heart failure (n=146), or Parkinson’s disease (n=2) were excluded for the restricted-complete cases sensitivity analysis (n=273) to minimize potential confounding from distinct clinical conditions. All participants provided written informed consent, and the research protocol was approved by Institutional Review Boards of all participating institutions.

**Figure 1.**
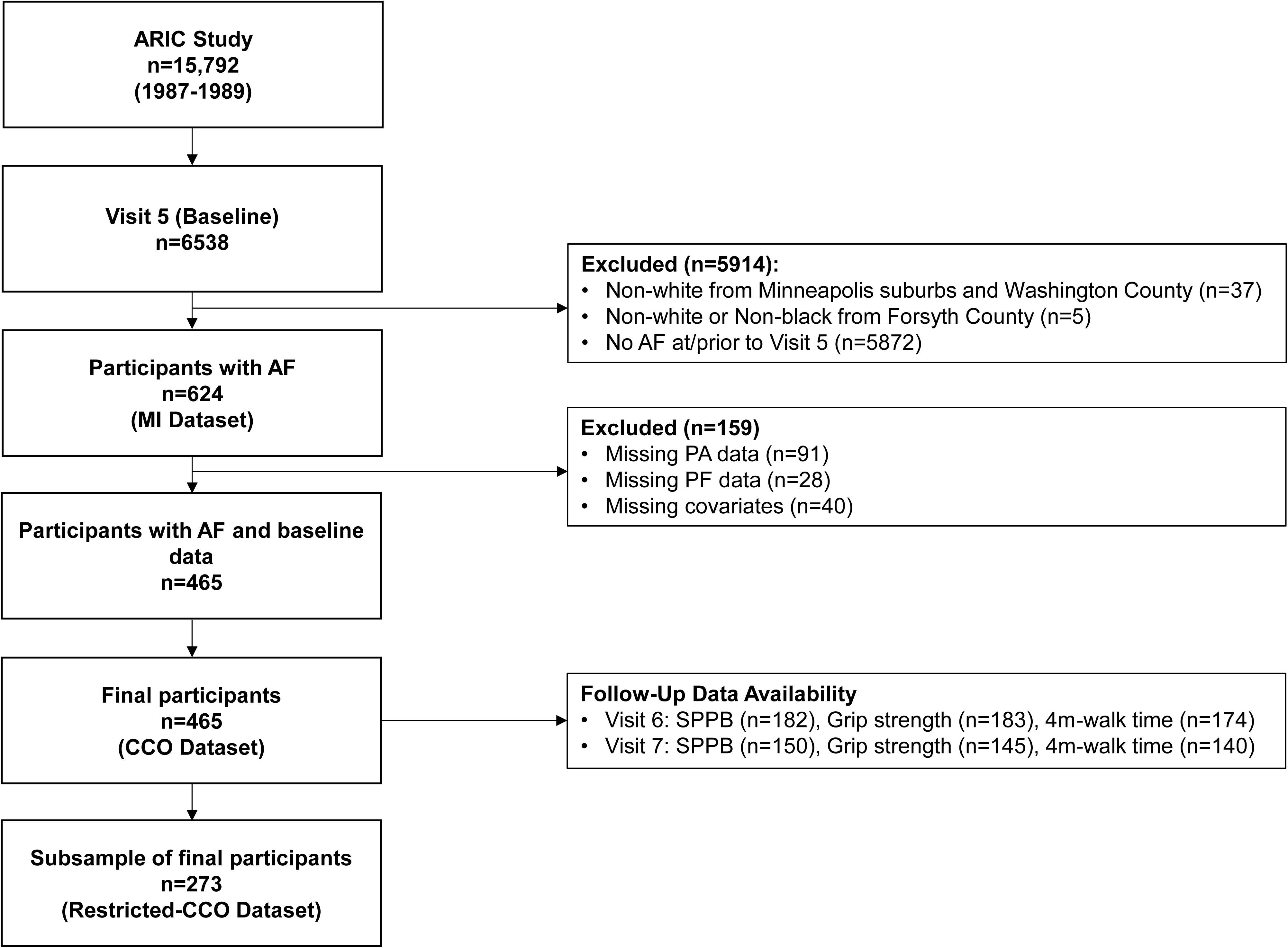
Flowchart of the selection of study participants. ARIC, atherosclerosis risk in communities. AF, atrial fibrillation; MI, multiple imputation; PA, physical activity; PF, physical function; CCO, complete case only; SPPB, short physical performance battery.

### Physical Activity

Physical activity was measured at Visit 5 using the modified Baecke Physical Activity Questionnaire, as previously applied in ARIC.^16, 17, 18^ Briefly, the questionnaire captures 3 physical activity components over the past year: work; leisure time sport; and leisure time non-sport. As most participants were retired at baseline, the work component was omitted due to age of participants who engaged little in formal work activities. The questionnaire was used to derive continuous component scores ranging from 1 to 5, with higher scores indicating greater physical activity. In addition, Metabolic Equivalent of Task (MET)-based estimates of physical activity were calculated from participants’ reported duration and frequency of the most common leisure time physical activities. These data were used to calculate the time and intensity of each physical activity, which were then summed across activities to calculate total weekly light and moderate-to-vigorous physical activity levels (MET-minutes/week).

### Physical Function

Physical function was assessed with the Short Physical Performance Battery and grip strength at Visits 5, 6 and 7, using similar protocols across visits.

#### Short Physical Performance Battery (SPPB)

Lower extremity physical function was measured using the SPPB,^19^ which includes 3 components: 4-meter walk test; standing balance; and repeated chair stands. The 3 components are each scored from 0 to 4, with the total SPPB score representing the sum of all 3 components. Thus, the SPPB score ranges from 0 to 12, with higher scores indicating better physical function. The 4-meter walk component was assessed by participants completing two trials at their usual walking pace. The faster result was used in the analysis. The 4-meter walk component was scored from 0 to 4 based on established time thresholds for completing 4-meters: 0 (unable to walk 4-meters); 1 (>8.70 seconds); 2 (≥6.21 to ≤8.70 seconds); 3 (≥4.82 to <6.21 seconds); and 4 (<4.82 seconds).

The standing balance component was evaluated through a balance test consisting of side-by-side, semi-tandem, and tandem stands, each performed for up to 10 seconds. The standing balance component was scored as a 1 if the participant held the side-by-side standing position for 10 seconds but could not hold a semi-tandem position for 10 seconds, a score of 2 if they could hold a semi-tandem position for 10 seconds but could not hold a full tandem position for more than 2 seconds, a score of 3 for holding the full tandem position for 3 to 9 seconds, and a score of 4 for holding the full tandem position for 10 seconds.

The chair stand component was measured through a chair stand test, which required participants to perform 5 chair stands using a chair with arms folded across the chest. The chair stand component was scored from 0 to 4 based time to complete all 5 chair stands: 0 (unable to finish in 60 seconds); 1 (≥16.7 to <60 seconds); 2 (≥13.7 to <16.7 seconds); 3 (≥11.2 to <13.7 seconds); and 4 (<11.2 seconds).

#### Grip Strength

Upper extremity physical function was evaluated by measuring grip strength (kg) using a Jamar dynamometer using a standardized protocol. Each participant’s grip strength was measured twice using their dominant hand, and the better result was recorded.^20, 21, 22^

#### 4-Meter Walk Test and Speed

To evaluate continuous performance data, the time taken to complete the 4-meter walk from the SPPB walk component was analyzed separately as a continuous variable. To facilitate comparison with literature reporting speed, 4-meter walk speed, calculated as distance divided by time, was also evaluated as a secondary outcome in supplemental analyses.^23^ Participants who were unable to complete the walk test were assigned a score of 0 within the composite SPPB analysis but were excluded from the continuous 4-meter walk time and speed analyses as a completion time or speed could not be determined.

### Covariates

Covariates included age, sex, race, center, education, body mass index (BMI), alcohol drinking, smoking, total cholesterol, low-density lipoprotein cholesterol (LDL), high-density lipoprotein cholesterol (HDL), triglyceride, hypertension, diabetes, history of stroke, history of heart failure, history of coronary heart disease, Mini-Mental State Examination (MMSE), and Center for Epidemiological Studies (CES) depression score. Information on sex, race, center, and education were self-reported at Visit 1. Race and center were combined in a single variable with 5 levels: Minneapolis-White, Washington-White, Forsyth-White, Forsyth-Black, Jackson-Black. Education was categorized into 6 levels: grade school (0-8 years); high school (no degree); high school graduate; vocational school; college (≥ 1 year of college, regardless of degree completion); and graduate/professional school. Data for other covariates were collected at Visit 5. BMI was calculated as weight (kg) divided by the square of height (m^2^). Alcohol drinking and smoking status were self-reported and categorized as current, former, never, or unknown. Blood samples collected at the visit were used to measure total cholesterol, LDL, HDL, and triglyceride using standardized protocols. Hypertension was defined as systolic blood pressure ≥ 140 mmHg, diastolic blood pressure ≥ 90 mmHg, or use of medication for high blood pressure. Diabetes was defined as fasting glucose level ≥ 126 mg/dL, non-fasting glucose level ≥ 200 mg/dL, use of medication for diabetes, or self-reported diagnosis of diabetes by a physician. History of stroke, heart failure, and coronary heart disease were defined based on the ARIC criteria.^24, 25, 26^ Cognitive function was assessed using MMSE scores,^27, 28^ and depression symptoms were evaluated using CES depression score.^20, 29^

### Statistical Analysis

Baseline characteristics were summarized for all participants and by tertiles of leisure time sport-related physical activity score (ranging from 1 to 5), serving as the primary representative metric of habitual physical activity. Continuous variables were reported as mean (SD), and categorical variables were reported as count (%). Linear mixed models were used to assess the association between physical activity and change in physical function, with Visits 5, 6, and 7 serving as the 3 timepoints. Fixed effects included time, physical activity, age, sex, race-center, education, BMI, alcohol drinking, smoking, total cholesterol, LDL, HDL, triglyceride, hypertension, diabetes, history of stroke, history of heart failure, history of coronary heart disease, MMSE, CES depression score, and time interactions for all variables. Subject ID was incorporated as a random intercept. To capture nonlinear changes in physical function over follow-up, time was treated as a categorical variable with 3 levels (Visits 5, 6, and 7). Model 1 was a crude model. Model 2 additionally included age, sex, and race-center as fixed effects, with ID as a random intercept. Model 3 further included education, BMI, alcohol drinking, smoking, total cholesterol, LDL, HDL, triglyceride, hypertension, diabetes, history of stroke, history of heart failure, history of coronary heart disease, MMSE, and CES depression score as fixed effects, with ID as a random intercept. Baseline cross-sectional associations between physical activity and physical function were assessed using linear regression models. Covariate adjustment followed the same approach described for the longitudinal analyses (Models 1, 2, and 3), excluding the random intercept for subject ID and all time-related parameters. For the restricted-complete cases sensitivity analysis, history of stroke and heart failure were omitted from Model 3.

All measures of physical activity were tested separately as continuous variables. For visualization of changes in physical function over follow-up, fully adjusted predicted means of physical function were plotted by tertiles of leisure time sport-related physical activity score, serving as the primary representative metric of habitual physical activity. MET-based estimates of total light and moderate-to-vigorous physical activity were divided by 100 to represent per 100 MET-minutes/week increase. Two-sided p-value < 0.05 was considered statistically significant.

Multiple imputation was used as part of the primary analysis to address missing baseline and follow-up data due to attrition, with additional sensitivity analyses using complete cases. The number and percentage of missingness across variables prior to imputation are delineated (Table S1). Multiple Imputation by Chained Equations (MICE) was used.^30^ Missing baseline data and follow-up physical function data were imputed using 20 iterations. As the complete cases data were suspected to be biased due to selective survival inherent in an older cohort with AF, continuous variables were imputed using linear regression without predictive mean matching to avoid restricting imputed values to the observed range of the healthier surviving sample.^31^ Results from MICE were pooled using Rubin’s rule to derive pooled parameter estimates.^30, 32^ All statistical analyses were performed using SAS software (Version 9.4; SAS Institute, Cary, NC, US).

## Results

### General Characteristics

Of the 624 participants with AF included in the primary analysis, the mean age was 78.5 ± 5.4 years; 52.6% were male, and 13.8% were Black participants (Table 1). The 624 participants with AF were divided into tertiles of leisure time sport-related physical activity score. Compared with the middle and low tertiles, participants in the highest tertile of physical activity were more likely to be male and White and had a lower prevalence of lower education, never smokers, never drinkers, hypertension, diabetes, stroke, heart failure, and coronary heart disease. They also had lower BMI, CES-D depression scores, and 4-meter walk times, as well as higher physical activity levels, SPPB scores, and grip strength.

**Table 1.** Baseline Characteristics Overall and by Tertiles of Leisure Time Sport-Related Physical Activity Score in the Multiple Imputation Data, ARIC cohort, 2011-2013.

|  |  | **Total**<br>**(n=624)** | **Low Tertile**<br>**(< 2.0)**<br>**(n=196)** | **Middle**<br>**Tertile**<br>**(2.0-2.7)**<br>**(n=204)** | **High**<br>**Tertile**<br>**(> 2.7)**<br>**(n=224)** |
| --- | --- | --- | --- | --- | --- |
| Age, years |  | 78.5 (5.4) | 78.6 (5.5) | 78.6 (5.5) | 78.2 (5.3) |
| Male, n (%) |  | 328 (52.6) | 75 (38.3) | 107 (52.6) | 146 (65.0) |
| Race, n (%) |  |  |  |  |  |
| White |  | 538 (86.2) | 167 (85.0) | 172 (84.6) | 199 (88.7) |
| Black |  | 86 (13.8) | 29 (15.0) | 32 (15.4) | 25 (11.3) |
| Education, n (%) |  |  |  |  |  |
| Grade school or 0 yrs |  | 31 (5.0) | 11 (5.6) | 12 (5.6) | 8 (3.9) |
| High school, no degree |  | 77 (12.3) | 26 (13.1) | 26 (12.9) | 25 (11.2) |
| High school graduate |  | 224 (35.9) | 76 (38.9) | 77 (37.8) | 71 (31.5) |
| Vocational school |  | 47 (7.5) | 15 (7.7) | 14 (6.9) | 18 (7.9) |
| College |  | 175 (28.0) | 51 (26.0) | 56 (27.5) | 68 (30.3) |
| Graduate/professional school |  | 70 (11.2) | 17 (8.8) | 19 (9.3) | 34 (15.1) |
| Smoking status, n (%) |  |  |  |  |  |
| Current smoker |  | 31 (5.0) | 8 (4.3) | 13 (6.1) | 10 (4.5) |
| Former smoker |  | 325 (52.1) | 97 (49.6) | 100 (49.2) | 128 (57.1) |
| Never smoker |  | 197 (31.6) | 71 (35.9) | 66 (32.6) | 60 (26.8) |
| Unknown |  | 71 (11.4) | 20 (10.2) | 25 (12.2) | 26 (11.6) |
| Drinking status, n (%) |  |  |  |  |  |
| Current drinker |  | 305 (48.9) | 91 (46.6) | 88 (43.0) | 126 (56.1) |
| Former drinker |  | 199 (31.9) | 65 (33.3) | 77 (37.9) | 57 (25.3) |
| Never drinker |  | 120 (19.2) | 40 (20.1) | 39 (19.1) | 41 (18.5) |
| BMI, kg/m<sup>2</sup> |  | 29.1 (6.1) | 30.6 (7.2) | 29.3 (6.1) | 27.7 (4.7) |
| Total cholesterol, mmol/L |  | 4.4 (1.0) | 4.3 (1.0) | 4.3 (1.1) | 4.4 (1.0) |
| LDL cholesterol, mmol/L |  | 2.4 (0.8) | 2.4 (0.8) | 2.4 (0.8) | 2.5 (0.9) |
| HDL cholesterol, mmol/L |  | 1.3 (0.4) | 1.3 (0.3) | 1.3 (0.4) | 1.3 (0.3) |
| Triglyceride, mmol/L | mean (SD) | 1.4 (0.8) | 1.4 (0.6) | 1.4 (0.9) | 1.4 (0.8) |
|  | median (25<sup>th</sup>-75<sup>th</sup> pctl) | 1.2 (0.9-1.6) | 1.3 (1.0-1.5) | 1.2 (0.9-1.6) | 1.2 (0.9-1.6) |
| Hypertension, n (%) |  | 490 (78.5) | 163 (83.1) | 164 (80.9) | 163 (72.5) |
| Diabetes, n (%) |  | 264 (42.3) | 99 (50.3) | 94 (46.4) | 71 (31.5) |
| Stroke history, n (%) |  | 66 (10.6) | 26 (13.4) | 25 (12.3) | 15 (6.5) |
| Heart failure history, n (%) |  | 241 (38.6) | 95 (48.4) | 77 (38.0) | 69 (30.6) |
| CHD history, n (%) |  | 200 (32.1) | 63 (31.9) | 70 (34.3) | 67 (30.0) |
| MMSE, score | mean (SD) | 26.4 (3.8) | 26.3 (4.0) | 26.3 (4.0) | 26.6 (3.5) |
|  | median (25<sup>th</sup>-75<sup>th</sup> pctl) | 27.8 (25.0-29.0) | 27.4 (25.0-29.0) | 27.8 (25.0-29.0) | 27.7 (25.3-29.0) |
| CES-D, score | mean (SD) | 3.6 (3.3) | 4.1 (3.4) | 3.8 (3.4) | 3.1 (3.0) |
|  | median (25<sup>th</sup>-75<sup>th</sup> pctl) | 3.0 (1.0-5.0) | 3.4 (1.0-6.3) | 3.0 (1.0-5.9) | 2.0 (1.0-4.2) |
| LTS physical activity, score |  | 2.4 (0.8) | 1.5 (0.3) | 2.3 (0.2) | 3.2 (0.5) |
| LTPA excluding sport, score |  | 2.2 (0.6) | 1.8 (0.5) | 2.1 (0.6) | 2.5 (0.6) |
| LSPA, MET-min/week | mean (SD) | 21.1 (90.2) | 2.7 (44.6) | 28.9 (106.0) | 30.3 (101.3) |
|  | median (25<sup>th</sup>-75<sup>th</sup> pctl) | 0.0 (0.0-0.0) | 0.0 (0.0-0.0) | 0.0 (0.0-1.3) | 0.0 (0.0-0.0) |
| MVSPA, MET-min/week | mean (SD) | 587.0 (749.7) | 66.0 (379.4) | 482.0 (584.3) | 1140.7 (758.9) |
|  | median (25<sup>th</sup>-75<sup>th</sup> pctl) | 406.9 (0.0-1008.5) | 0.0 (0.0-4.8) | 366.9 (0.2-801.1) | 1051.5 (616.6-1606.7) |
| SPPB, score | mean (SD) | 7.8 (3.1) | 6.8 (3.3) | 7.6 (3.0) | 8.9 (2.7) |
|  | median (25<sup>th</sup>-75<sup>th</sup> pctl) | 8.0 (6.0-10.0) | 7.2 (4.2-9.9) | 8.0 (5.9-10.0) | 9.0 (7.1-11.0) |
| 4-meter walk time, seconds |  | 5.3 (2.0) | 6.1 (2.6) | 5.3 (1.7) | 4.6 (1.4) |
| Grip strength, kg |  | 28.2 (10.5) | 24.6 (9.2) | 28.1 (9.9) | 31.6 (10.9) |

Data are presented as n (%) for categorical variables and mean (SD) for continuous variables unless otherwise stated.
BMI, body mass index; LDL, low-density lipoprotein cholesterol; HDL, high-density lipoprotein cholesterol; CHD, coronary heart disease; MMSE, mini-mental state examination; CES-D, center for epidemiological studies-depression; LTS, leisure time sport-related; LTPA, leisure time physical activity; LSPA, total light sport physical activity; MVSPA, total moderate-to-vigorous sport physical activity; SPPB, short physical performance battery (total score).
Unit conversion: to convert cholesterol to milligrams per deciliter, multiply by 38.67; to convert triglycerides to milligrams per deciliter, multiply by 88.57.

Baseline characteristics for the multiple imputation dataset and complete cases dataset were similar (Table S2). Compared with the complete cases dataset, the multiple imputation dataset had a higher prevalence of diabetes, lower SPPB and grip strength, and longer 4-meter walk times.

### Change in Physical Function

The median follow-up time from Visit 5 to 6 was 4.9 years and 6.6 years from Visit 5 to 7. The unadjusted mean SPPB (Visit 5: 7.8 ± 3.1, Visit 6: 6.3 ± 3.8, Visit 7: 5.7 ± 3.8) and grip strength (Visit 5: 28.2 ± 10.5, Visit 6: 26.0 ± 10.6, Visit 7: 23.8 ± 10.1) decreased over time, while 4-meter walk time increased over time (Visit 5: 5.3 ± 2.0, Visit 6: 5.6 ± 1.9, Visit 7: 5.8 ± 2.3) (Figure 2). The unadjusted mean 4-meter walk speed decreased over time (Visit 5: 0.830 ± 0.241, Visit 6: 0.772 ± 0.272, Visit 7: 0.770 ± 0.272) (Figure S1). Among complete cases, there were modest declines in SPPB (Visit 5: 8.4 ± 2.7, Visit 6: 7.8 ± 2.9, Visit 7: 7.4 ± 3.1) and grip strength (Visit 5: 29.2 ± 10.2, Visit 6: 27.7 ± 9.8, Visit 7: 27.1 ± 9.2), and minimal changes in 4-meter walk time (Visit 5: 5.0 ± 1.5, Visit 6: 4.9 ± 1.5, Visit 7: 4.9 ± 1.7) over follow-up (Figure 2). Small increases in 4-meter walk speed were observed among complete cases (Visit 5: 0.868 ± 0.219, Visit 6: 0.879 ± 0.221, Visit 7: 0.884 ± 0.209) over follow-up (Figure S1).

**Figure 2.**
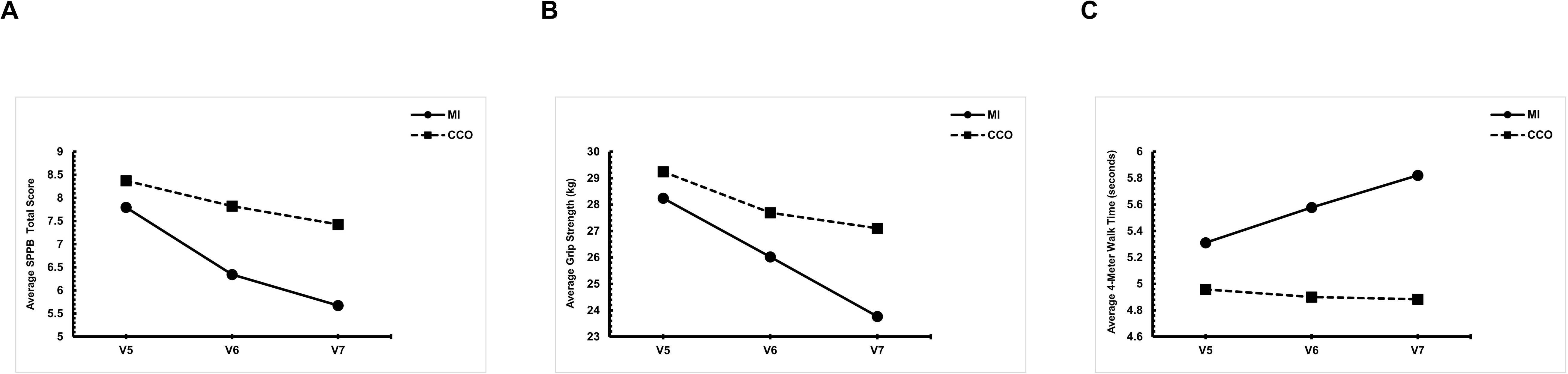
Average changes in physical function during follow-up by multiple imputation (n=624) and complete case only (n=465) datasets: (A) SPPB; (B) Grip Strength; (C) 4-Meter Walk Time. MI, multiple imputation; CCO, complete case only; SPPB, short physical performance battery; V5, visit 5; V6, visit 6; V7, visit 7.

### Cross-Sectional Association between Physical Activity and Physical Function

In fully adjusted models with SPPB as the outcome, greater leisure time sport-related physical activity score (β: 0.83, 95% CI: 0.50 to 1.17), leisure time non-sport physical activity score (β: 0.68, 95% CI: 0.28 to 1.08), and total moderate-to-vigorous sport physical activity (β: 0.07, 95% CI: 0.04 to 0.11) were significantly associated with higher SPPB scores at baseline (Table 2).

**Table 2.** Baseline Cross-Sectional Associations between Measures of Physical Activity and Physical Function in the Multiple Imputation Data (n=624)

| | Model | Short Physical Performance Battery<br>$\beta$ (95% CI) | Grip Strength<br>$\beta$ (95% CI) | 4-Meter Walk Time<br>$\beta$ (95% CI) |
| --- | --- | --- | --- | --- |
| LTS physical activity, score | Model 1<sup>a</sup> | **1.25 (0.87, 1.63)** | **4.17 (2.99, 5.35)** | **-0.81 (-1.05, -0.56)** |
|  | Model 2<sup>b</sup> | **1.09 (0.72, 1.46)** | **2.51 (1.63, 3.38)** | **-0.67 (-0.92, -0.43)** |
|  | Model 3<sup>c</sup> | **0.83 (0.50, 1.17)** | **2.37 (1.47, 3.26)** | **-0.43 (-0.64, -0.21)** |
| LTPA excluding sport, score | Model 1<sup>a</sup> | **1.16 (0.71, 1.60)** | **3.05 (1.59, 4.50)** | **-0.76 (-1.03, -0.48)** |
|  | Model 2<sup>b</sup> | **0.99 (0.57, 1.41)** | **1.42 (0.40, 2.45)** | **-0.58 (-0.84, -0.32)** |
|  | Model 3<sup>c</sup> | **0.68 (0.28, 1.08)** | **1.17 (0.09, 2.24)** | **-0.27 (-0.52, -0.03)** |
| LSPA, 100 MET-min/week | Model 1<sup>a</sup> | 0.14 (-0.18, 0.46) | -0.92 (-1.88, 0.04) | -0.17 (-0.37, 0.04) |
|  | Model 2<sup>b</sup> | 0.18 (-0.12, 0.48) | 0.07 (-0.62, 0.76) | -0.19 (-0.38, 0.00) |
|  | Model 3<sup>c</sup> | 0.14 (-0.12, 0.40) | 0.07 (-0.62, 0.76) | -0.16 (-0.33, 0.01) |
| MVSPA, 100 MET-min/week | Model 1<sup>a</sup> | **0.12 (0.09, 0.16)** | **0.36 (0.24, 0.48)** | **-0.08 (-0.10, -0.05)** |
|  | Model 2<sup>b</sup> | **0.10 (0.06, 0.14)** | **0.21 (0.12, 0.30)** | **-0.05 (-0.08, -0.03)** |
|  | Model 3<sup>c</sup> | **0.07 (0.04, 0.11)** | **0.19 (0.10, 0.29)** | **-0.03 (-0.05, -0.01)** |

Each measure of physical activity was tested separately.
CI, confidence interval; LTS, leisure time sport-related; LTPA, leisure time physical activity; LSPA, total light sport physical activity; MVSPA, total moderate-to-vigorous sport physical activity.
<sup>a</sup> Crude model.
<sup>b</sup> Adjusted for age, sex, and race-center.
<sup>c</sup> Adjusted for age, sex, race-center, education, BMI, drinking status, smoking status, total cholesterol, LDL cholesterol, HDL cholesterol, triglyceride, hypertension, diabetes, history of stroke, history of heart failure, history of coronary heart disease, MMSE, and CES-D.

Similarly, greater leisure time sport-related physical activity score (β: 2.37, 95% CI: 1.47 to 3.26), leisure time non-sport physical activity score (β: 1.17, 95% CI: 0.09 to 2.24), and total moderate-to-vigorous sport physical activity (β: 0.19, 95% CI: 0.10 to 0.29) were significantly associated with higher grip strength at baseline (Table 2).

Greater leisure time sport-related physical activity score (β: -0.43, 95% CI: -0.64 to -0.21), leisure time non-sport physical activity score (β: -0.27, 95% CI: -0.52 to -0.03), and total moderate-to-vigorous sport physical activity (β: -0.03, 95% CI: -0.05 to -0.01) were significantly associated with shorter 4-meter walk time at baseline (Table 2). Similar results were observed when evaluating 4-meter walk speed as the outcome (Table S3). Sensitivity analyses consisting of complete cases and restricted-complete cases yielded similar results for SPPB, grip strength, and 4-meter walk time (Tables S4-S5).

### Association between Physical Activity and Change in Physical Function

Regarding SPPB, the only significant interaction term was the leisure time non-sport physical activity and time interaction for Visit 6 vs. Visit 5 (β: -0.58, 95% CI: -1.04 to -0.12). All other physical activity and time interaction terms were not significant (Table 3). Similar results were found from the sensitivity analyses consisting of complete cases (Table S6) and restricted-complete cases (Table S7).

**Table 3.** Association between Measures of Physical Activity and Change in Short Physical Performance Battery Total Score in the Multiple Imputation Data (n=624)

|  | Model | V6 vs. V5 | V7 vs. V5 |
| --- | --- | --- | --- |
| LTS physical activity, score | Model 1<sup>a</sup> | 0.13 (-0.41, 0.67) | 0.20 (-0.32, 0.72) |
|  | Model 2<sup>b</sup> | 0.06 (-0.47, 0.59) | 0.09 (-0.40, 0.57) |
|  | Model 3<sup>c</sup> | -0.11 (-0.60, 0.39) | -0.11 (-0.62, 0.39) |
| LTPA excluding sport, score | Model 1<sup>a</sup> | -0.34 (-0.88, 0.19) | -0.30 (-1.10, 0.49) |
|  | Model 2<sup>b</sup> | -0.33 (-0.83, 0.18) | -0.29 (-1.08, 0.49) |
|  | Model 3<sup>c</sup> | **-0.58 (-1.04, -0.12)** | -0.68 (-1.48, 0.13) |
| LSPA, 100 MET-min/week | Model 1<sup>a</sup> | 0.25 (-0.14, 0.63) | 0.24 (-0.18, 0.66) |
|  | Model 2<sup>b</sup> | 0.20 (-0.19, 0.60) | 0.15 (-0.30, 0.60) |
|  | Model 3<sup>c</sup> | 0.14 (-0.21, 0.49) | 0.17 (-0.21, 0.54) |
| MVSPA, 100 MET-min/week | Model 1<sup>a</sup> | 0.01 (-0.03, 0.05) | 0.02 (-0.03, 0.06) |
|  | Model 2<sup>b</sup> | 0.00 (-0.04, 0.05) | 0.00 (-0.05, 0.04) |
|  | Model 3<sup>c</sup> | -0.02 (-0.07, 0.03) | -0.04 (-0.09, 0.01) |

Each measure of physical activity was tested separately.
CI, confidence interval; LTS, leisure time sport-related; LTPA, leisure time physical activity; LSPA, total light sport physical activity; MVSPA, total moderate-to-vigorous sport physical activity.
<sup>a</sup> Crude model with subject ID as random intercept.
<sup>b</sup> Adjusted for age, sex, and race-center, with subject ID as random intercept.
<sup>c</sup> Adjusted for age, sex, race-center, education, BMI, drinking status, smoking status, total cholesterol, LDL cholesterol, HDL cholesterol, triglyceride, hypertension, diabetes, history of stroke, history of heart failure, history of coronary heart disease, MMSE, and CES-D, with subject ID as random intercept.

When assessing grip strength as the outcome, all physical activity and time interaction terms were not significant (Table 4). Sensitivity analyses restricted to complete cases (Table S8) and restricted-complete cases (Table S9) yielded similar results.

**Table 4.** Association between Measures of Physical Activity and Change in Grip Strength in the Multiple Imputation Data (n=624)

|  | Model | V6 vs. V5 | V7 vs. V5 |
| --- | --- | --- | --- |
| LTS physical activity, score | Model 1<sup>a</sup> | -0.20 (-1.56, 1.15) | -0.05 (-1.46, 1.36) |
|  | Model 2<sup>b</sup> | 0.04 (-1.20, 1.27) | 0.02 (-1.38, 1.41) |
|  | Model 3<sup>c</sup> | -0.02 (-1.36, 1.33) | -0.34 (-1.57, 0.89) |
| LTPA excluding sport, score | Model 1<sup>a</sup> | 0.12 (-1.33, 1.57) | -1.20 (-2.91, 0.52) |
|  | Model 2<sup>b</sup> | 0.29 (-1.08, 1.65) | -0.97 (-2.62, 0.69) |
|  | Model 3<sup>c</sup> | 0.42 (-1.46, 2.30) | -1.50 (-3.27, 0.26) |
| LSPA, 100 MET-min/week | Model 1<sup>a</sup> | -0.01 (-0.97, 0.95) | 0.71 (-0.23, 1.65) |
|  | Model 2<sup>b</sup> | -0.14 (-1.15, 0.88) | 0.56 (-0.38, 1.50) |
|  | Model 3<sup>c</sup> | -0.23 (-1.22, 0.75) | 0.62 (-0.21, 1.44) |
| MVSPA, 100 MET-min/week | Model 1<sup>a</sup> | -0.06 (-0.19, 0.06) | -0.05 (-0.18, 0.09) |
|  | Model 2<sup>b</sup> | -0.05 (-0.16, 0.07) | -0.06 (-0.19, 0.07) |
|  | Model 3<sup>c</sup> | -0.06 (-0.18, 0.05) | -0.10 (-0.21, 0.01) |

Each measure of physical activity was tested separately.
CI, confidence interval; LTS, leisure time sport-related; LTPA, leisure time physical activity; LSPA, total light sport physical activity; MVSPA, total moderate-to-vigorous sport physical activity.
<sup>a</sup> Crude model with subject ID as random intercept.
<sup>b</sup> Adjusted for age, sex, and race-center, with subject ID as random intercept.
<sup>c</sup> Adjusted for age, sex, race-center, education, BMI, drinking status, smoking status, total cholesterol, LDL cholesterol, HDL cholesterol, triglyceride, hypertension, diabetes, history of stroke, history of heart failure, history of coronary heart disease, MMSE, and CES-D, with subject ID as random intercept.

Similarly, when assessing 4-meter walk time as the outcome, there were no significant physical activity and time interactions (Table 5), with consistent trends observed across both complete cases (Table S10) and restricted-complete cases sensitivity analyses (Table S11). Similar results were observed when evaluating 4-meter walk speed as the outcome (Table S12).

**Table 5.** Association between Measures of Physical Activity and Change in 4-Meter Walk Time in the Multiple Imputation Data (n=624)

|  | Model | V6 vs. V5 | V7 vs. V5 |
| --- | --- | --- | --- |
| LTS physical activity, score | Model 1<sup>a</sup> | 0.02 (-0.23, 0.28) | -0.15 (-0.51, 0.22) |
|  | Model 2<sup>b</sup> | 0.04 (-0.21, 0.29) | -0.10 (-0.47, 0.28) |
|  | Model 3<sup>c</sup> | 0.04 (-0.23, 0.30) | -0.11 (-0.52, 0.30) |
| LTPA excluding sport, score | Model 1<sup>a</sup> | 0.31 (-0.01, 0.62) | 0.02 (-0.46, 0.49) |
|  | Model 2<sup>b</sup> | 0.23 (-0.06, 0.51) | 0.01 (-0.46, 0.48) |
|  | Model 3<sup>c</sup> | 0.23 (-0.12, 0.57) | 0.06 (-0.44, 0.57) |
| LSPA, 100 MET-min/week | Model 1<sup>a</sup> | 0.01 (-0.19, 0.21) | 0.08 (-0.16, 0.33) |
|  | Model 2<sup>b</sup> | 0.03 (-0.17, 0.23) | 0.08 (-0.17, 0.34) |
|  | Model 3<sup>c</sup> | 0.01 (-0.18, 0.20) | 0.07 (-0.15, 0.29) |
| MVSPA, 100 MET-min/week | Model 1<sup>a</sup> | 0.00 (-0.03, 0.02) | -0.02 (-0.05, 0.01) |
|  | Model 2<sup>b</sup> | -0.01 (-0.03, 0.02) | -0.01 (-0.05, 0.02) |
|  | Model 3<sup>c</sup> | -0.01 (-0.03, 0.02) | -0.01 (-0.05, 0.03) |

Each measure of physical activity was tested separately.
CI, confidence interval; LTS, leisure time sport-related; LTPA, leisure time physical activity; LSPA, total light sport physical activity; MVSPA, total moderate-to-vigorous sport physical activity.
<sup>a</sup> Crude model with subject ID as random intercept.
<sup>b</sup> Adjusted for age, sex, and race-center, with subject ID as random intercept.
<sup>c</sup> Adjusted for age, sex, race-center, education, BMI, drinking status, smoking status, total cholesterol, LDL cholesterol, HDL cholesterol, triglyceride, hypertension, diabetes, history of stroke, history of heart failure, history of coronary heart disease, MMSE, and CES-D, with subject ID as random intercept.

Fully adjusted predicted means by tertiles of leisure time sport-related physical activity score showed decreases in SPPB and grip strength and increases in 4-meter walk time over follow-up. Similar patterns across the 3 tertile groups for all physical function measures were observed (Figure 3; Figure S2).

**Figure 3.**
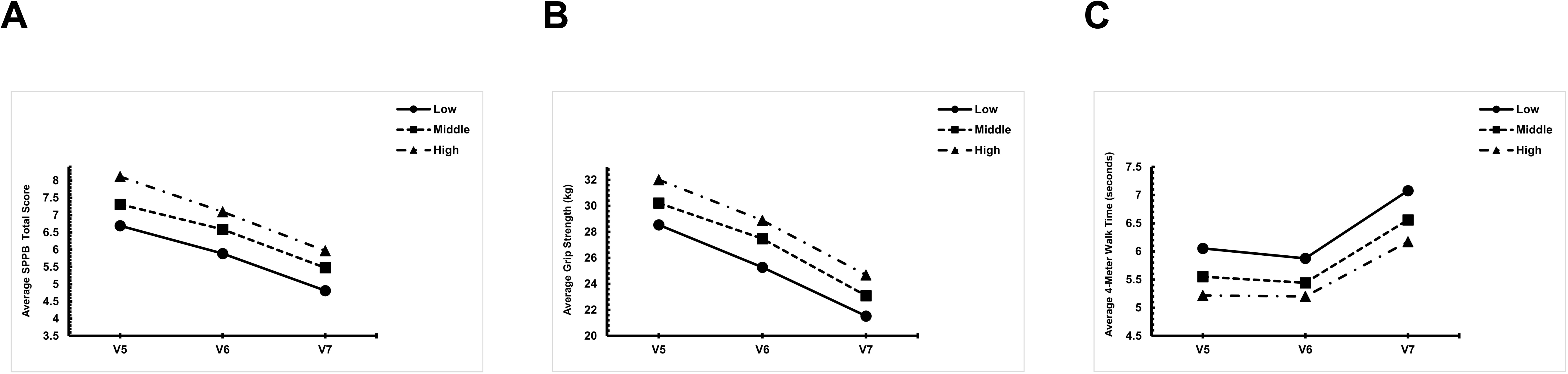
Fully adjusted predicted means of physical function by tertiles of leisure time sport-related physical activity score over follow-up, estimated from linear mixed models including time interactions (n=624): (A) SPPB; (B) Grip Strength; (C) 4-Meter Walk Time. SPPB, short physical performance battery; V5, visit 5; V6, visit 6; V7, visit 7.

## Discussion

In this prospective community-based cohort study investigating the association between habitual physical activity and change in physical function in participants with AF, leisure time sport-related physical activity score, leisure time non-sport physical activity score, and total moderate-to-vigorous sport physical activity had significant baseline cross-sectional associations with SPPB, grip strength, and 4-meter walk time where greater habitual physical activity levels were associated with higher SPPB, grip strength, and shorter 4-meter walk time at baseline. However, greater habitual physical activity levels were not significantly associated with slower declines in SPPB, grip strength, and slower increases in 4-meter walk time among participants with AF over follow-up. These findings are partially consistent with prior studies, which have reported significant effects of physical activity on physical function in randomized controlled trials consisting of small samples of AF patients and short follow-up periods.^14^ Specifically, the cross-sectional associations of habitual physical activity and physical function observed in this study are consistent. In contrast, there was a lack of evidence of significant longitudinal associations in this study. While prior studies have demonstrated that structured exercise interventions may improve physical function in individuals with AF, the current study did not observe slower declines in physical function over long-term follow-up from greater habitual physical activity. Such contrast could be attributed to differences in the physical activity measures and amount of physical activity. As prior studies implemented structured exercise programs in the context of randomized controlled trials, the overall volume and intensity of physical activity may be greater than that of the habitual physical activity measures used in the current study. This difference may explain the lack of significant longitudinal associations in the current study. While the current study observed a significant association between leisure time non-sport physical activity and change in SPPB, indicating that greater leisure time non-sport physical activity at baseline was associated with a faster decline in SPPB, this isolated finding should be interpreted cautiously and may represent a chance observation considering the number of statistical comparisons performed. Importantly, given the current study’s sample size and the parameter estimates for longitudinal associations being near zero across all 3 physical function measures, the findings may represent a lack of clinically meaningful longitudinal associations rather than an underpowered statistical artifact. The current study’s strengths of having a large sample size, an extensive follow-up period, and using objectively measured physical function measures add valuable information to the evidence base. Our results uniquely highlight that greater habitual physical activity may not be associated with slower worsening of physical function over time in AF participants, and the beneficial effects of short-term structured exercise programs on physical function in randomized controlled trials consisting of AF patients may not translate to habitual physical activity in everyday life. Nevertheless, because greater habitual physical activity was associated with better baseline physical function but not with future trajectories, AF participants with greater habitual physical activity may maintain better physical function over time.

Despite the null longitudinal findings, this study has several key clinical implications. As physical function measures such as SPPB, grip strength, and 4-meter walk time may represent the underlying state of metabolic dysfunction, systemic inflammation, oxidative stress, and neurodegeneration,^33, 34, 35^ it is plausible that changes in such multi-system aspects are slow to respond to habitual physical activity unless the activity is targeted.^36^ This may partly explain the lack of significant longitudinal associations in the current study. Furthermore, the current study’s results suggest that the intensity of physical activity may play an important role. Given the lack of significant longitudinal associations with change in physical function measures for leisure time sport-related physical activity score and total moderate-to-vigorous sport physical activity (MET-minutes/week), higher intensity physical activities may be necessary to observe clinically meaningful changes in SPPB, grip strength, and 4-meter walk time among participants with AF.^37^ The low intensity of habitual physical activity may contribute to the lack of significant longitudinal associations between physical activity and change in physical function. Among people with AF, achieving and maintaining physical activity at sufficient intensity levels may be particularly challenging due to AF-related symptoms such as fatigue, dyspnea, palpitations, and exercise intolerance, which may limit participation in higher-intensity physical activities that are more likely to induce physiological adaptations and improvements in physical function.^38^ Furthermore, compromised cardiac output associated with AF may constrain exercise capacity, which in turn may reduce exercise performance.^38^ Thus, future studies should aim to evaluate safe and effective intensity levels for AF patients, considering the impact of symptom burden and exercise capacity. The relatively older age of AF participants in the current study compared to prior studies suggests that the timing of initiation of physical activity may also be important. Initiation of physical activity earlier in the disease course or prior to substantial functional decline may be necessary to achieve meaningful long-term benefits in physical function.^39, 40^ Considering the mean age of 78.5 ± 5.4 years for the participants in the current study, it is reasonable that the advanced age limits functional plasticity due to biological aging processes and thus lack of significant longitudinal associations between physical activity and change in physical function.^41^ In the presence of AF, the combination of AF-specific symptom burden, reduced exercise capacity, and age-related functional decline may attenuate the relationship between habitual physical activity and changes in physical function, thereby contributing to the lack of significant longitudinal associations observed in this study.

The current study has several strengths including the large sample size, extensive follow-up period, multiple measures of physical activity and objectively measured physical function parameters, and incorporation of multiple covariates. Although the study offers notable strengths, certain limitations should be considered. The measures of physical activity in this study were self-reported, implying the possibility of measurement error. However, the validation of measures of physical activity may reduce such errors.^42^ Furthermore, participants with complete follow-up were likely healthier than those who were unable or unwilling to attend visits, introducing potential selection bias due to visit non-attendance. While multiple imputation analyses produced similar results to the sensitivity analyses, the possibility of selection bias related to visit non-attendance cannot be excluded. The relatively older age of participants in the current study compared to prior studies suggests that generalizability of the findings to younger AF populations may be limited. Despite the incorporation of multiple covariates, the possibility of residual confounding persists. Finally, the observed cross-sectional associations may also reflect reverse causation, whereby participants with better physical function are more likely to remain physically active.

## Conclusion

The findings of this study demonstrated that greater habitual physical activity levels were associated with higher SPPB, grip strength, and shorter 4-meter walk time at baseline in participants with AF. However, greater habitual physical activity levels were not significantly associated with slower declines in SPPB, grip strength, and slower increases in 4-meter walk time in participants with AF. Whether structured exercise programs or higher-intensity physical activity can improve physical functioning in individuals with AF requires future randomized intervention studies. These findings emphasize the potential importance of the timing of physical activity initiation, and disease stage, as well as the limited physiological adaptability associated with advanced age among individuals with AF. Future studies should aim to develop targeted exercise interventions with sufficient sample size, follow-up period, sufficient dose of physical activity, objectively measured physical function, and appropriate intervention timings for individuals with AF.

## Data Availability

Data are available from the Atherosclerosis Risk in Communities (ARIC) Study upon approval of a data use application; restrictions apply and data are not publicly available.

## Acknowledgements

The authors thank the staff and participants of the ARIC study for their important contributions.

## Supplemental Material

Tables S1-S12, Figures S1-S2

