## Supplemental Material for "Association of Physical Activity with Change in Physical Function in Individuals with Atrial Fibrillation: The Atherosclerosis Risk in Communities (ARIC) Study"

**Table S1.** Frequency and Percentage of Missing Data Prior to Multiple Imputation (n=624)

|  | **Number of Missing** | **% of Missing** |
| --- | --- | --- |
| MMSE | 6 | 0.96 |
| CHD history | 7 | 1.12 |
| Hypertension | 18 | 2.88 |
| Total cholesterol | 20 | 3.21 |
| HDL cholesterol | 20 | 3.21 |
| Triglyceride | 20 | 3.21 |
| CES-D | 23 | 3.69 |
| LDL cholesterol | 25 | 4.01 |
| Smoking status | 38 | 6.09 |
| Diabetes | 39 | 6.25 |
| Drinking status | 67 | 10.74 |
| BMI | 72 | 11.54 |
| LSPA | 84 | 13.46 |
| MVSPA | 84 | 13.46 |
| LTPA excluding sport | 86 | 13.78 |
| LTS physical activity | 91 | 14.58 |
| SPPB | 97 | 15.54 |
| Grip strength | 99 | 15.87 |
| 4-meter walk time | 110 | 17.63 |
| Visit 6 Grip strength | 427 | 68.43 |
| Visit 6 SPPB | 428 | 68.59 |
| Visit 6 4-meter walk time | 436 | 69.87 |
| Visit 7 SPPB | 459 | 73.56 |
| Visit 7 Grip strength | 465 | 74.52 |
| Visit 7 4-meter walk time | 470 | 75.32 |

MMSE, mini-mental state examination; CHD, coronary heart disease; HDL, high-density lipoprotein cholesterol; CES-D, center for epidemiological studies-depression; LDL, low-density lipoprotein cholesterol; BMI, body mass index; LSPA, total light sport physical activity; MVSPA, total moderate-to-vigorous sport physical activity; LTS, leisure time sport-related; LTPA, leisure time physical activity; SPPB, short physical performance battery (total score). **Table S2**. Comparison of Baseline Characteristics between MI and CCO Data, ARIC cohort, 2011- 2013

|  | | **MI Data**  **(n=624)** | **CCO Data**  **(n=465)** |
| --- | --- | --- | --- |
| Age, years | | 78.5 (5.4) | 77.8 (5.3) |
| Male, n (%) | | 328 (52.6) | 258 (55.5) |
| Race, n (%) | |  |  |
| White | | 538 (86.2) | 413 (88.8) |
| Black | | 86 (13.8) | 52 (11.2) |
| Education, n (%) | |  |  |
| Grade school or 0 yrs | | 31 (5.0) | 16 (3.4) |
| High school, no degree | | 77 (12.3) | 47 (10.1) |
| High school graduate | | 224 (35.9) | 166 (35.7) |
| Vocational school | | 47 (7.5) | 40 (8.6) |
| College | | 175 (28.0) | 140 (30.1) |
| Graduate/professional school | | 70 (11.2) | 56 (12.0) |
| Smoking status, n (%) | |  |  |
| Current smoker | | 31 (5.0) | 21 (4.5) |
| Former smoker | | 325 (52.1) | 266 (57.2) |
| Never smoker | | 197 (31.6) | 150 (32.3) |
| Unknown | | 71 (11.4) | 28 (6.0) |
| Drinking status, n (%) | |  |  |
| Current drinker | | 305 (48.9) | 244 (52.5) |
| Former drinker | | 199 (31.9) | 135 (29.0) |
| Never drinker | | 120 (19.2) | 86 (18.5) |
| BMI, kg/m^2^ | | 29.1 (6.1) | 29.2 (5.9) |
| Total cholesterol, mmol/L | | 4.4 (1.0) | 4.3 (1.0) |
| LDL cholesterol, mmol/L | | 2.4 (0.8) | 2.4 (0.9) |
| HDL cholesterol, mmol/L | | 1.3 (0.4) | 1.3 (0.3) |
| Triglyceride, mmol/L | mean (SD) | 1.4 (0.8) | 1.4 (0.7) |
|  | median (25^th^-75^th^ pctl) | 1.2 (0.9-1.6) | 1.2 (0.9-1.6) |
| Hypertension, n (%) | | 490 (78.5) | 365 (78.5) |
| Diabetes, n (%) | | 264 (42.3) | 171 (36.8) |
| Stroke history, n (%) | | 66 (10.6) | 44 (9.5) |
| Heart failure history, n (%) | | 241 (38.6) | 168 (36.1) |
| CHD history, n (%) | | 200 (32.1) | 141 (30.3) |
| MMSE, score | mean (SD) | 26.4 (3.8) | 27.2 (2.7) |
|  | median (25^th^-75^th^ pctl) | 27.8 (25.0-29.0) | 28.0 (26.0-29.0) |
| CES-D, score | mean (SD) | 3.6 (3.3) | 3.4 (3.2) |
|  | median (25^th^-75^th^ pctl) | 3.0 (1.0-5.0) | 3.0 (1.0-5.0) |
| LTS physical activity, score | | 2.4 (0.8) | 2.5 (0.8) |
| LTPA excluding sport, score | | 2.2 (0.6) | 2.2 (0.6) |

| LSPA, MET-min/week | mean (SD) | 21.1 (90.2) | 22.6 (90.3) |
| --- | --- | --- | --- |
|  | median (25^th^-75^th^ pctl) | 0.0 (0.0-0.0) | 0.0 (0.0-0.0) |
| MVSPA, MET-min/week | mean (SD) | 587.0 (749.7) | 645.1 (751.4) |
|  | median (25^th^-75^th^ pctl) | 406.9 (0.0-1008.5) | 455.6 (0.0-1063.0) |
| SPPB, score | mean (SD) | 7.8 (3.1) | 8.4 (2.7) |
|  | median (25^th^-75^th^ pctl) | 8.0 (6.0-10.0) | 9.0 (6.0-11.0) |
| 4-meter walk time, seconds | | 5.3 (2.0) | 5.0 (1.5) |
| Grip strength, kg | | 28.2 (10.5) | 29.2 (10.2) |

Data are presented as n (%) for categorical variables and mean (SD) for continuous variables unless otherwise stated.

MI, multiple imputation; CCO, complete case only; BMI, body mass index; LDL, low-density lipoprotein cholesterol; HDL, high-density lipoprotein cholesterol; CHD, coronary heart disease; MMSE, mini-mental state examination; CES-D, center for epidemiological studies-depression; LTS, leisure time sport-related; LTPA, leisure time physical activity; LSPA, total light sport physical activity; MVSPA, total moderate-to-vigorous sport physical activity; SPPB, short physical performance battery (total score).

Unit conversion: to convert cholesterol to milligrams per deciliter, multiply by 38.67; to convert triglycerides to milligrams per deciliter, multiply by 88.57.
**Table S3**. Baseline Cross-Sectional Associations between Measures of Physical Activity and 4-Meter Walk Speed in the Multiple Imputation Data (n=624)

|  | Model | 4-Meter Walk Speed  β (95% CI) |
| --- | --- | --- |
| LTS physical activity, score | Model 1^a^ | **0.118 (0.092, 0.144)** |
|  | Model 2^b^ | **0.106 (0.081, 0.130)** |
|  | Model 3^c^ | **0.075 (0.051, 0.099)** |
| LTPA excluding sport, score | Model 1^a^ | **0.125 (0.092, 0.158)** |
|  | Model 2^b^ | **0.101 (0.071, 0.130)** |
|  | Model 3^c^ | **0.062 (0.034, 0.090)** |
| LSPA, 100 MET-min/week | Model 1^a^ | 0.021 (-0.002, 0.044) |
|  | Model 2^b^ | **0.024 (0.003, 0.044)** |
|  | Model 3^c^ | **0.021 (0.003, 0.039)** |
| MVSPA, 100 MET-min/week | Model 1^a^ | **0.012 (0.010, 0.015)** |
|  | Model 2^b^ | **0.010 (0.007, 0.013)** |
|  | Model 3^c^ | **0.007 (0.004, 0.009)** |

Each measure of physical activity was tested separately.

CI, confidence interval; LTS, leisure time sport-related; LTPA, leisure time physical activity; LSPA, total light sport physical activity; MVSPA, total moderate-to-vigorous sport physical activity.

^a^ Crude model.

^b^ Adjusted for age, sex, and race-center.

^c^ Adjusted for age, sex, race-center, education, BMI, drinking status, smoking status, total cholesterol, LDL cholesterol, HDL cholesterol, triglyceride, hypertension, diabetes, history of stroke, history of heart failure, history of coronary heart disease, MMSE, and CES-D.
**Table S4**. Baseline Cross-Sectional Associations between Measures of Physical Activity and Physical Function among Complete Cases (n=465)

|  | Model | Short Physical Performance Battery  β (95% CI) | Grip Strength  β (95% CI) | 4-Meter Walk Time  β (95% CI) |
| --- | --- | --- | --- | --- |
| LTS physical activity, score | Model 1^a^ | **1.03 (0.72, 1.34)** | **3.72 (2.56, 4.89)** | **-0.63 (-0.80, -0.46)** |
|  | Model 2^b^ | **0.95 (0.64, 1.26)** | **2.39 (1.50, 3.27)** | **-0.57 (-0.73, -0.40)** |
|  | Model 3^c^ | **0.72 (0.41, 1.04)** | **2.22 (1.28, 3.17)** | **-0.35 (-0.51, -0.18)** |
| LTPA excluding sport, score | Model 1^a^ | **1.02 (0.64, 1.40)** | **2.99 (1.55, 4.42)** | **-0.65 (-0.86, -0.45)** |
|  | Model 2^b^ | **0.90 (0.53, 1.27)** | **1.47 (0.40, 2.55)** | **-0.52 (-0.73, -0.32)** |
|  | Model 3^c^ | **0.63 (0.25, 1.01)** | 1.15 (-0.01, 2.31) | **-0.26 (-0.46, -0.06)** |
| LSPA, 100 MET-min/week | Model 1^a^ | 0.03 (-0.25, 0.31) | -0.94 (-1.97, 0.09) | -0.09 (-0.24, 0.07) |
|  | Model 2^b^ | 0.09 (-0.18, 0.36) | 0.13 (-0.63, 0.89) | -0.11 (-0.26, 0.03) |
|  | Model 3^c^ | 0.08 (-0.18, 0.33) | 0.18 (-0.59, 0.95) | -0.11 (-0.25, 0.02) |
| MVSPA, 100 MET-min/week | Model 1^a^ | **0.11 (0.08, 0.14)** | **0.35 (0.23, 0.47)** | **-0.06 (-0.08, -0.05)** |
|  | Model 2^b^ | **0.09 (0.06, 0.12)** | **0.20 (0.11, 0.29)** | **-0.05 (-0.07, -0.03)** |
|  | Model 3^c^ | **0.07 (0.03, 0.10)** | **0.18 (0.08, 0.28)** | **-0.03 (-0.04, -0.01)** |

Each measure of physical activity was tested separately.

CI, confidence interval; LTS, leisure time sport-related; LTPA, leisure time physical activity; LSPA, total light sport physical activity; MVSPA, total moderate-to-vigorous sport physical activity.

^a^ Crude model.

^b^ Adjusted for age, sex, and race-center.

^c^ Adjusted for age, sex, race-center, education, BMI, drinking status, smoking status, total cholesterol, LDL cholesterol, HDL cholesterol, triglyceride, hypertension, diabetes, history of stroke, history of heart failure, history of coronary heart disease, MMSE, and CES-D.

**Table S5**. Baseline Cross-Sectional Associations between Measures of Physical Activity and Physical Function among Complete Cases Without Stroke, Heart Failure, or Parkinson’s Disease (n=273)

|  | Model | Short Physical Performance Battery  β (95% CI) | Grip Strength  β (95% CI) | 4-Meter Walk Time  β (95% CI) |
| --- | --- | --- | --- | --- |
| LTS physical activity, score | Model 1^a^ | **0.81 (0.43, 1.19)** | **4.32 (2.84, 5.80)** | **-0.44 (-0.62, -0.27)** |
|  | Model 2^b^ | **0.68 (0.30, 1.06)** | **2.84 (1.79, 3.89)** | **-0.38 (-0.55, -0.20)** |
|  | Model 3^c^ | **0.68 (0.27, 1.08)** | **2.87 (1.75, 3.99)** | **-0.26 (-0.43, -0.09)** |
| LTPA excluding sport, score | Model 1^a^ | **0.72 (0.26, 1.18)** | **3.20 (1.36, 5.05)** | **-0.51 (-0.72, -0.30)** |
|  | Model 2^b^ | **0.66 (0.21, 1.11)** | **1.95 (0.68, 3.23)** | **-0.41 (-0.61, -0.21)** |
|  | Model 3^c^ | **0.67 (0.18, 1.16)** | **1.81 (0.42, 3.21)** | **-0.29 (-0.50, -0.08)** |
| LSPA, 100 MET-min/week | Model 1^a^ | 0.03 (-0.28, 0.34) | -0.42 (-1.66, 0.82) | -0.13 (-0.27, 0.01) |
|  | Model 2^b^ | 0.07 (-0.23, 0.37) | 0.22 (-0.62, 1.06) | **-0.14 (-0.27, 0.00)** |
|  | Model 3^c^ | 0.09 (-0.21, 0.40) | 0.32 (-0.54, 1.18) | **-0.15 (-0.28, -0.03)** |
| MVSPA, 100 MET-min/week | Model 1^a^ | **0.08 (0.04, 0.11)** | **0.35 (0.21, 0.49)** | **-0.04 (-0.06, -0.03)** |
|  | Model 2^b^ | **0.06 (0.03, 0.10)** | **0.19 (0.09, 0.30)** | **-0.03 (-0.05, -0.02)** |
|  | Model 3^c^ | **0.06 (0.02, 0.10)** | **0.19 (0.09, 0.30)** | **-0.02 (-0.04, -0.01)** |

Each measure of physical activity was tested separately.

CI, confidence interval; LTS, leisure time sport-related; LTPA, leisure time physical activity; LSPA, total light sport physical activity; MVSPA, total moderate-to-vigorous sport physical activity.

^a^ Crude model.

^b^ Adjusted for age, sex, and race-center.

^c^ Adjusted for age, sex, race-center, education, BMI, drinking status, smoking status, total cholesterol, LDL cholesterol, HDL cholesterol, triglyceride, hypertension, diabetes, history of coronary heart disease, MMSE, and CES-D.

**Table S6**. Association between Measures of Physical Activity and Change in Short Physical Performance Battery Total Score among Complete Cases (n=465)

|  | Model | V6 vs. V5 | V7 vs. V5 |
| --- | --- | --- | --- |
| LTS physical activity, score | Model 1^a^ | 0.15 (-0.25, 0.56) | 0.26 (-0.17, 0.69) |
|  | Model 2^b^ | 0.07 (-0.34, 0.48) | 0.12 (-0.31, 0.55) |
|  | Model 3^c^ | -0.03 (-0.49, 0.43) | 0.06 (-0.42, 0.54) |
| LTPA excluding sport, score | Model 1^a^ | -0.35 (-0.87, 0.18) | -0.08 (-0.65, 0.49) |
|  | Model 2^b^ | -0.33 (-0.85, 0.19) | -0.12 (-0.69, 0.44) |
|  | Model 3^c^ | **-0.68 (-1.26, -0.11)** | -0.50 (-1.12, 0.11) |
| LSPA, 100 MET-min/week | Model 1^a^ | 0.11 (-0.18, 0.41) | 0.25 (-0.08, 0.58) |
|  | Model 2^b^ | 0.09 (-0.19, 0.38) | 0.18 (-0.14, 0.51) |
|  | Model 3^c^ | 0.10 (-0.19, 0.40) | 0.21 (-0.13, 0.55) |
| MVSPA, 100 MET-min/week | Model 1^a^ | 0.00 (-0.04, 0.04) | 0.00 (-0.04, 0.04) |
|  | Model 2^b^ | -0.01 (-0.05, 0.03) | -0.01 (-0.06, 0.03) |
|  | Model 3^c^ | -0.03 (-0.07, 0.01) | -0.04 (-0.09, 0.00) |

Each measure of physical activity was tested separately.

CI, confidence interval; LTS, leisure time sport-related; LTPA, leisure time physical activity; LSPA, total light sport physical activity; MVSPA, total moderate-to-vigorous sport physical activity.

^a^ Crude model with subject ID as random intercept.

^b^ Adjusted for age, sex, and race-center, with subject ID as random intercept.

^c^ Adjusted for age, sex, race-center, education, BMI, drinking status, smoking status, total cholesterol, LDL cholesterol, HDL cholesterol, triglyceride, hypertension, diabetes, history of stroke, history of heart failure, history of coronary heart disease, MMSE, and CES-D, with subject ID as random intercept.

**Table S7**. Association between Measures of Physical Activity and Change in Short Physical Performance Battery Total Score among Complete Cases Without Stroke, Heart Failure, or Parkinson’s Disease (n=273)

|  | Model | V6 vs. V5 | V7 vs. V5 |
| --- | --- | --- | --- |
| LTS physical activity, score | Model 1^a^ | 0.07 (-0.42, 0.57) | 0.30 (-0.22, 0.82) |
|  | Model 2^b^ | -0.05 (-0.55, 0.45) | 0.10 (-0.42, 0.62) |
|  | Model 3^c^ | -0.44 (-1.03, 0.15) | -0.03 (-0.60, 0.55) |
| LTPA excluding sport, score | Model 1^a^ | -0.14 (-0.77, 0.49) | 0.04 (-0.64, 0.71) |
|  | Model 2^b^ | -0.17 (-0.78, 0.45) | -0.01 (-0.67, 0.65) |
|  | Model 3^c^ | **-0.75 (-1.45, -0.05)** | -0.52 (-1.27, 0.24) |
| LSPA, 100 MET-min/week | Model 1^a^ | 0.09 (-0.24, 0.43) | 0.35 (-0.04, 0.73) |
|  | Model 2^b^ | 0.09 (-0.23, 0.42) | 0.26 (-0.12, 0.63) |
|  | Model 3^c^ | 0.11 (-0.23, 0.44) | 0.34 (-0.05, 0.73) |
| MVSPA, 100 MET-min/week | Model 1^a^ | -0.01 (-0.05, 0.04) | 0.00 (-0.04, 0.05) |
|  | Model 2^b^ | -0.03 (-0.07, 0.02) | -0.03 (-0.07, 0.02) |
|  | Model 3^c^ | **-0.07 (-0.12, -0.02)** | **-0.06 (-0.11, -0.01)** |

Each measure of physical activity was tested separately.

CI, confidence interval; LTS, leisure time sport-related; LTPA, leisure time physical activity; LSPA, total light sport physical activity; MVSPA, total moderate-to-vigorous sport physical activity.

^a^ Crude model with subject ID as random intercept.

^b^ Adjusted for age, sex, and race-center, with subject ID as random intercept.

^c^ Adjusted for age, sex, race-center, education, BMI, drinking status, smoking status, total cholesterol, LDL cholesterol, HDL cholesterol, triglyceride, hypertension, diabetes, history of coronary heart disease, MMSE, and CES-D, with subject ID as random intercept.

**Table S8**. Association between Measures of Physical Activity and Change in Grip Strength among Complete Cases (n=465)

|  | Model | V6 vs. V5 | V7 vs. V5 |
| --- | --- | --- | --- |
| LTS physical activity, score | Model 1^a^ | -0.76 (-1.83, 0.31) | -0.19 (-1.34, 0.95) |
|  | Model 2^b^ | -0.61 (-1.68, 0.45) | 0.01 (-1.13, 1.15) |
|  | Model 3^c^ | -0.19 (-1.40, 1.02) | -0.20 (-1.48, 1.08) |
| LTPA excluding sport, score | Model 1^a^ | -0.59 (-1.97, 0.79) | -1.23 (-2.76, 0.30) |
|  | Model 2^b^ | -0.61 (-1.96, 0.74) | -0.91 (-2.43, 0.60) |
|  | Model 3^c^ | -0.10 (-1.60, 1.40) | -1.36 (-3.03, 0.32) |
| LSPA, 100 MET-min/week | Model 1^a^ | -0.11 (-0.87, 0.65) | 0.54 (-0.33, 1.40) |
|  | Model 2^b^ | -0.23 (-0.98, 0.52) | 0.52 (-0.34, 1.38) |
|  | Model 3^c^ | -0.30 (-1.08, 0.49) | 0.52 (-0.38, 1.42) |
| MVSPA, 100 MET-min/week | Model 1^a^ | -0.07 (-0.17, 0.03) | -0.08 (-0.18, 0.02) |
|  | Model 2^b^ | -0.07 (-0.17, 0.03) | -0.07 (-0.18, 0.04) |
|  | Model 3^c^ | -0.06 (-0.17, 0.05) | -0.10 (-0.21, 0.02) |

Each measure of physical activity was tested separately.

CI, confidence interval; LTS, leisure time sport-related; LTPA, leisure time physical activity; LSPA, total light sport physical activity; MVSPA, total moderate-to-vigorous sport physical activity.

^a^ Crude model with subject ID as random intercept.

^b^ Adjusted for age, sex, and race-center, with subject ID as random intercept.

^c^ Adjusted for age, sex, race-center, education, BMI, drinking status, smoking status, total cholesterol, LDL cholesterol, HDL cholesterol, triglyceride, hypertension, diabetes, history of stroke, history of heart failure, history of coronary heart disease, MMSE, and CES-D, with subject ID as random intercept.

**Table S9**. Association between Measures of Physical Activity and Change in Grip Strength among Complete Cases Without Stroke, Heart Failure, or Parkinson’s Disease (n=273)

|  | Model | V6 vs. V5 | V7 vs. V5 |
| --- | --- | --- | --- |
| LTS physical activity, score | Model 1^a^ | -0.80 (-2.01, 0.40) | -0.68 (-1.97, 0.61) |
|  | Model 2^b^ | -0.64 (-1.85, 0.58) | -0.58 (-1.88, 0.71) |
|  | Model 3^c^ | -0.35 (-1.81, 1.11) | -0.85 (-2.28, 0.58) |
| LTPA excluding sport, score | Model 1^a^ | -0.29 (-1.82, 1.24) | -1.61 (-3.30, 0.08) |
|  | Model 2^b^ | -0.26 (-1.77, 1.24) | -1.47 (-3.14, 0.20) |
|  | Model 3^c^ | -0.36 (-2.06, 1.35) | **-2.53 (-4.43, -0.63)** |
| LSPA, 100 MET-min/week | Model 1^a^ | -0.36 (-1.17, 0.45) | 0.63 (-0.32, 1.57) |
|  | Model 2^b^ | -0.37 (-1.17, 0.43) | 0.58 (-0.35, 1.51) |
|  | Model 3^c^ | -0.27 (-1.12, 0.57) | 0.67 (-0.30, 1.64) |
| MVSPA, 100 MET-min/week | Model 1^a^ | -0.04 (-0.14, 0.07) | -0.11 (-0.22, 0.00) |
|  | Model 2^b^ | -0.03 (-0.14, 0.08) | **-0.12 (-0.24, 0.00)** |
|  | Model 3^c^ | -0.05 (-0.18, 0.07) | **-0.15 (-0.28, -0.03)** |

Each measure of physical activity was tested separately.

CI, confidence interval; LTS, leisure time sport-related; LTPA, leisure time physical activity; LSPA, total light sport physical activity; MVSPA, total moderate-to-vigorous sport physical activity.

^a^ Crude model with subject ID as random intercept.

^b^ Adjusted for age, sex, and race-center, with subject ID as random intercept.

^c^ Adjusted for age, sex, race-center, education, BMI, drinking status, smoking status, total cholesterol, LDL cholesterol, HDL cholesterol, triglyceride, hypertension, diabetes, history of coronary heart disease, MMSE, and CES-D, with subject ID as random intercept.

**Table S10**. Association between Measures of Physical Activity and Change in 4-Meter Walk Time among Complete Cases (n=465)

|  | Model | V6 vs. V5 | V7 vs. V5 |
| --- | --- | --- | --- |
| LTS physical activity, score | Model 1^a^ | -0.21 (-0.42, 0.00) | -0.13 (-0.35, 0.09) |
|  | Model 2^b^ | -0.13 (-0.34, 0.08) | -0.06 (-0.28, 0.15) |
|  | Model 3^c^ | -0.06 (-0.29, 0.17) | -0.08 (-0.32, 0.16) |
| LTPA excluding sport, score | Model 1^a^ | 0.05 (-0.23, 0.32) | 0.07 (-0.22, 0.37) |
|  | Model 2^b^ | -0.01 (-0.27, 0.26) | 0.08 (-0.21, 0.37) |
|  | Model 3^c^ | 0.12 (-0.17, 0.41) | 0.16 (-0.16, 0.47) |
| LSPA, 100 MET-min/week | Model 1^a^ | 0.03 (-0.12, 0.18) | -0.02 (-0.18, 0.15) |
|  | Model 2^b^ | 0.03 (-0.11, 0.17) | -0.04 (-0.20, 0.12) |
|  | Model 3^c^ | 0.00 (-0.14, 0.15) | -0.01 (-0.18, 0.16) |
| MVSPA, 100 MET-min/week | Model 1^a^ | -0.01 (-0.03, 0.01) | -0.01 (-0.03, 0.01) |
|  | Model 2^b^ | -0.01 (-0.03, 0.01) | 0.00 (-0.02, 0.02) |
|  | Model 3^c^ | 0.00 (-0.03, 0.02) | 0.01 (-0.02, 0.03) |

Each measure of physical activity was tested separately.

CI, confidence interval; LTS, leisure time sport-related; LTPA, leisure time physical activity; LSPA, total light sport physical activity; MVSPA, total moderate-to-vigorous sport physical activity.

^a^ Crude model with subject ID as random intercept.

^b^ Adjusted for age, sex, and race-center, with subject ID as random intercept.

^c^ Adjusted for age, sex, race-center, education, BMI, drinking status, smoking status, total cholesterol, LDL cholesterol, HDL cholesterol, triglyceride, hypertension, diabetes, history of stroke, history of heart failure, history of coronary heart disease, MMSE, and CES-D, with subject ID as random intercept.

**Table S11**. Association between Measures of Physical Activity and Change in 4-Meter Walk Time among Complete Cases Without Stroke, Heart Failure, or Parkinson’s Disease (n=273)

|  | Model | V6 vs. V5 | V7 vs. V5 |
| --- | --- | --- | --- |
| LTS physical activity, score | Model 1^a^ | **-0.26 (-0.49, -0.02)** | -0.12 (-0.37, 0.13) |
|  | Model 2^b^ | -0.17 (-0.41, 0.07) | -0.06 (-0.30, 0.19) |
|  | Model 3^c^ | 0.06 (-0.22, 0.34) | -0.08 (-0.35, 0.19) |
| LTPA excluding sport, score | Model 1^a^ | 0.04 (-0.26, 0.34) | 0.20 (-0.12, 0.52) |
|  | Model 2^b^ | 0.00 (-0.29, 0.29) | 0.21 (-0.10, 0.52) |
|  | Model 3^c^ | 0.20 (-0.13, 0.54) | 0.27 (-0.08, 0.62) |
| LSPA, 100 MET-min/week | Model 1^a^ | 0.09 (-0.07, 0.25) | -0.01 (-0.19, 0.17) |
|  | Model 2^b^ | 0.09 (-0.06, 0.24) | -0.01 (-0.19, 0.16) |
|  | Model 3^c^ | 0.08 (-0.08, 0.23) | 0.03 (-0.15, 0.20) |
| MVSPA, 100 MET-min/week | Model 1^a^ | -0.01 (-0.03, 0.01) | 0.00 (-0.03, 0.02) |
|  | Model 2^b^ | -0.01 (-0.03, 0.01) | 0.00 (-0.02, 0.02) |
|  | Model 3^c^ | 0.01 (-0.02, 0.03) | 0.02 (-0.01, 0.04) |

Each measure of physical activity was tested separately.

CI, confidence interval; LTS, leisure time sport-related; LTPA, leisure time physical activity; LSPA, total light sport physical activity; MVSPA, total moderate-to-vigorous sport physical activity.

^a^ Crude model with subject ID as random intercept.

^b^ Adjusted for age, sex, and race-center, with subject ID as random intercept.

^c^ Adjusted for age, sex, race-center, education, BMI, drinking status, smoking status, total cholesterol, LDL cholesterol, HDL cholesterol, triglyceride, hypertension, diabetes, history of coronary heart disease, MMSE, and CES-D, with subject ID as random intercept.
**Table S12**. Association between Measures of Physical Activity and Change in 4-Meter Walk Speed in the Multiple Imputation Data (n=624)

|  | Model | V6 vs. V5 | V7 vs. V5 |
| --- | --- | --- | --- |
| LTS physical activity, score | Model 1^a^ | -0.004 (-0.046, 0.037) | -0.013 (-0.069, 0.043) |
|  | Model 2^b^ | -0.015 (-0.054, 0.024) | -0.022 (-0.077, 0.033) |
|  | Model 3^c^ | -0.019 (-0.062, 0.024) | -0.023 (-0.080, 0.033) |
| LTPA excluding sport, score | Model 1^a^ | -0.032 (-0.074, 0.010) | -0.027 (-0.100, 0.046) |
|  | Model 2^b^ | -0.024 (-0.065, 0.017) | -0.021 (-0.091, 0.049) |
|  | Model 3^c^ | -0.032 (-0.075, 0.012) | -0.033 (-0.098, 0.031) |
| LSPA, 100 MET-min/week | Model 1^a^ | -0.003 (-0.031, 0.025) | -0.003 (-0.036, 0.031) |
|  | Model 2^b^ | -0.005 (-0.033, 0.023) | 0.000 (-0.033, 0.033) |
|  | Model 3^c^ | -0.002 (-0.028, 0.025) | 0.001 (-0.025, 0.027) |
| MVSPA, 100 MET-min/week | Model 1^a^ | 0.000 (-0.004, 0.004) | -0.001 (-0.006, 0.004) |
|  | Model 2^b^ | -0.001 (-0.005, 0.002) | -0.002 (-0.007, 0.002) |
|  | Model 3^c^ | -0.002 (-0.005, 0.002) | -0.003 (-0.007, 0.002) |

Each measure of physical activity was tested separately.

CI, confidence interval; LTS, leisure time sport-related; LTPA, leisure time physical activity; LSPA, total light sport physical activity; MVSPA, total moderate-to-vigorous sport physical activity.

^a^ Crude model with subject ID as random intercept.

^b^ Adjusted for age, sex, and race-center, with subject ID as random intercept.

^c^ Adjusted for age, sex, race-center, education, BMI, drinking status, smoking status, total cholesterol, LDL cholesterol, HDL cholesterol, triglyceride, hypertension, diabetes, history of stroke, history of heart failure, history of coronary heart disease, MMSE, and CES-D, with subject ID as random intercept.
**Figure S1.** Average changes in 4-meter walk speed during follow-up by multiple imputation (n=624) and complete case only (n=465) datasets. MI, multiple imputation; CCO, complete case only; V5, visit 5; V6, visit 6; V7, visit 7.
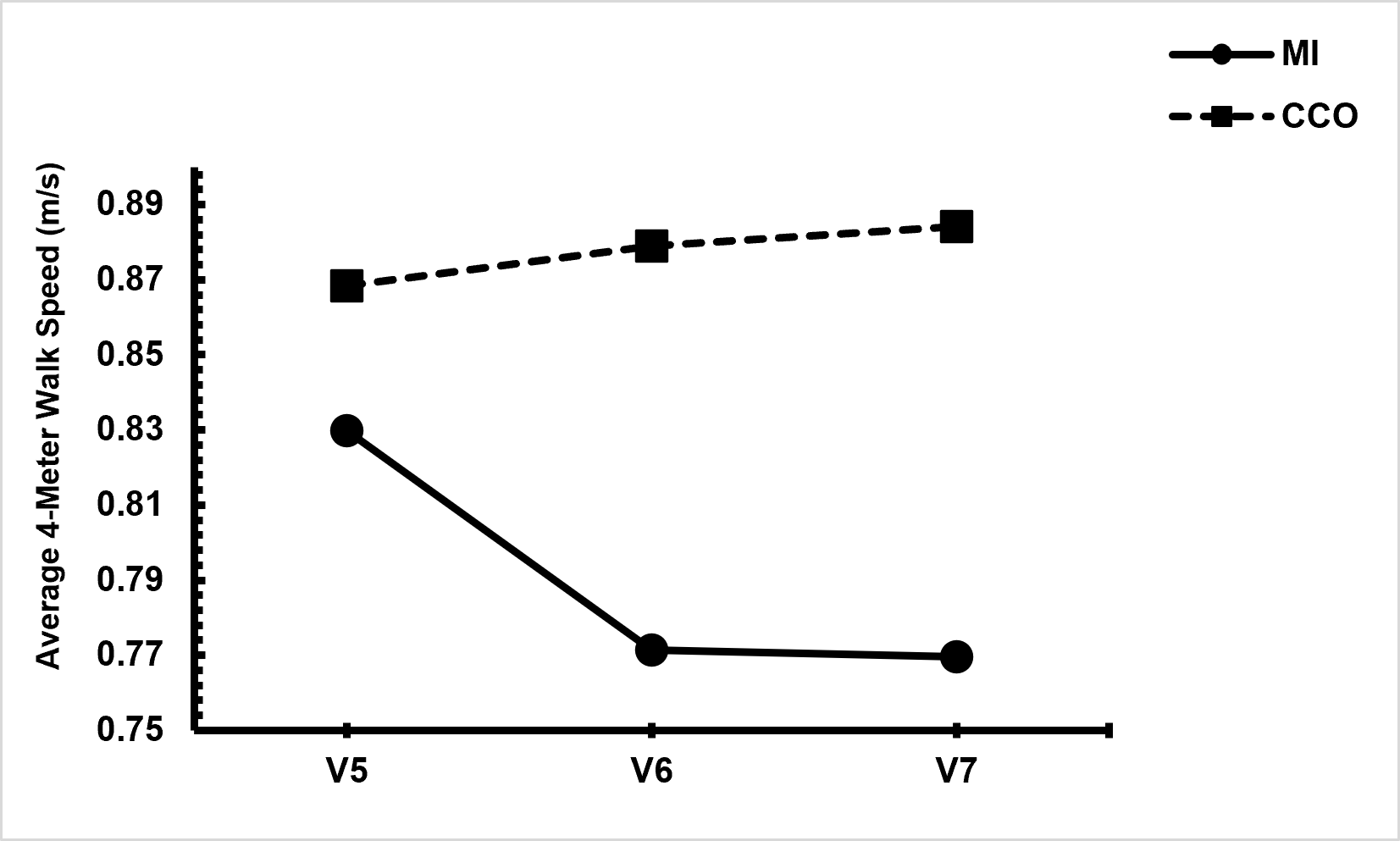

**Figure S2.** Fully adjusted predicted means of 4-meter walk speed by tertiles of leisure time sport-related physical activity score over follow-up, estimated from linear mixed models including time interactions (n=624). V5, visit 5; V6, visit 6; V7, visit 7.
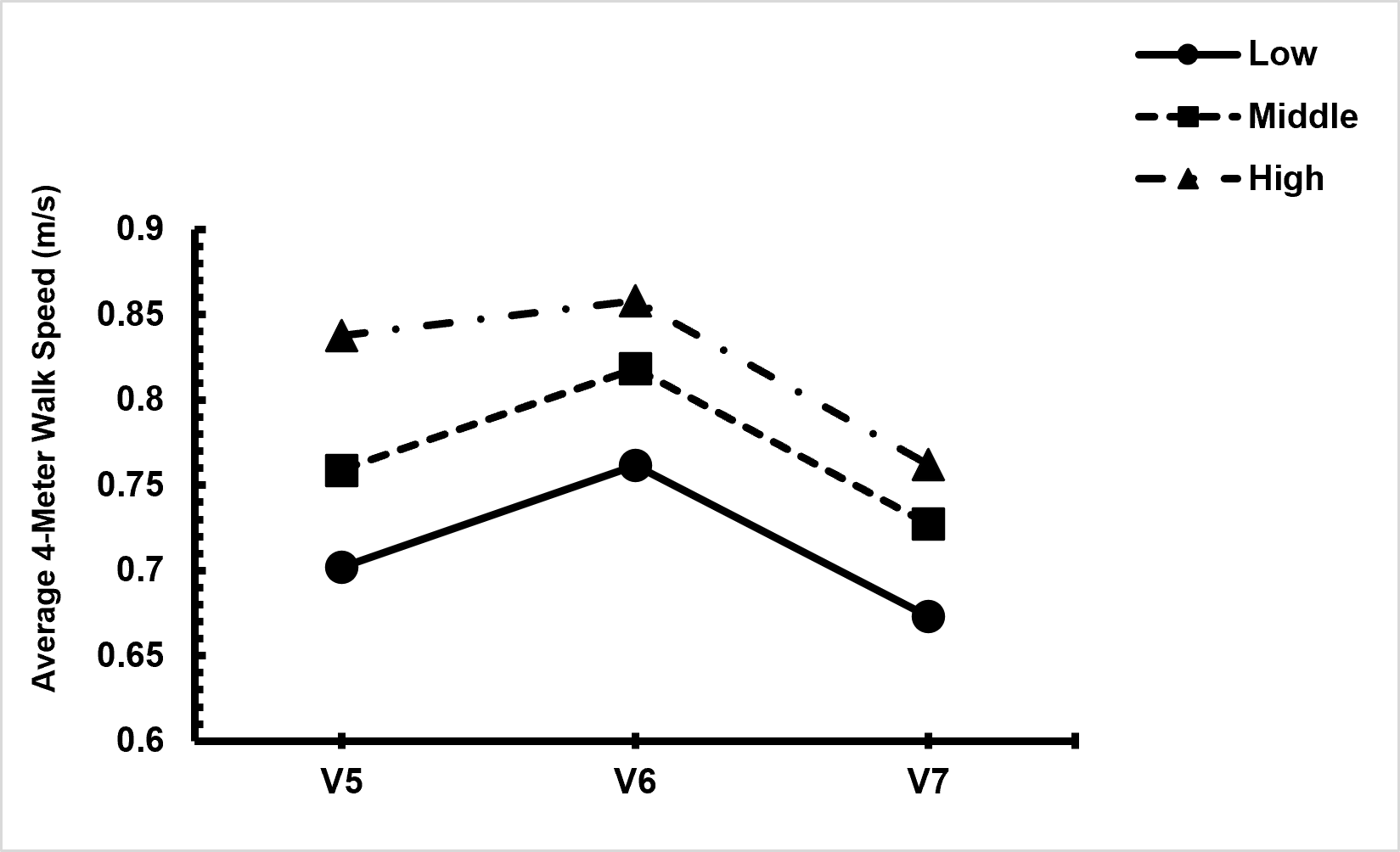
